# Molecular Investigation of DENV serotypes in the dengue outbreak of 2022 in Nepal

**DOI:** 10.1101/2023.05.26.23290534

**Authors:** Nishan Katuwal, Aastha Shrestha, Urusha Ranjitkar, Suraj Jakibanjar, Surendra Kumar Madhup, Dipesh Tamrakar, Rajeev Shrestha

## Abstract

**Introduction:** Dengue, a viral infection highly prevalent in tropical regions, exhibits local variations in risk that are influenced by a combination of climatic, socioeconomic, and environmental factors. The disease is caused by four distinct yet closely related serotypes of the dengue virus: DENV-1, DENV-2, DENV-3, and DENV-4.

**Objectives:** The objective of this study is to identify the different serotypes of dengue virus responsible for the 2022 outbreak in Nepal, where dengue has been prevalent since 2006 but with limited availability of molecular information on the serotypes.

**Methodology:** Serum samples from suspected dengue patients visiting Dhulikhel Hospital were analyzed using Dengue Ag and IgM/IgG Ab Kit test, for the presence of IgG/IgM antibodies or NS1 Ag. The positive samples were stored at -80 °C, and 89 samples were selected for further analysis. RNA was extracted from those positive samples using the Zymo Quick RNA Viral Kit, and RT-PCR was performed using the Sacace Dengue Real Genotype qPCR kit to identify the dengue virus serotypes present.

**Results:** The study included 89 samples, of which a higher percentage of sero-positivity was observed in females (52%) compared to males. Positive cases were distributed in 14 different districts, with the highest percentage (58.4%) in Kavre. Molecular investigation, of 53 out of 89 serologically positive samples, by qPCR revealed that DENV1 was the predominant serotype, followed by DEN3 (24.5%) and DENV2 (16.9%). DENV4 was not detected in any of the samples. The average Ct value of all serotypes was 17.6, with DENV3 having the lowest Ct value of 16.6, indicating a high viremia.

**Conclusion:** Our study, although limited in its coverage of Nepal, has provided molecular information on the serotypes responsible for the 2022 dengue outbreak. The high prevalence of DENV1 was observed, while prevalence of DENV3 was accompanied by high viral load. This information provided valuable insight into the circulating serotypes of the virus in the region.

## Introduction

Dengue Fever, caused by Dengue Virus (DENV), and transmitted by Aedes aegypti or Aedes albopictus, is a major cause of morbidity and mortality worldwide.^1^ The infection could vary from asymptomatic to acute febrile illness and to severe hemorrhagic fever/dengue shock syndromes (DHF/DSS).^2^ About 4 billion people are at risk of infection globally^1^ and the risk profile is increasing with climate change with spread to new geographical areas.^3-5^

At the local level, urbanization (especially unplanned), is associated with dengue transmission through multiple social and environmental factors including population density, human mobility, water storage practices.^6^ Consequently, disease risks change and shift with climate change in tropical and subtropical areas, and vectors might adapt to new environment and climate.^7^ Nepal is at higher risk, for dengue, due to its extensive altitudinal and climatic variations, with new infections being observed at higher altitude.^8-10^ In Nepal, dengue epidemic outbreaks occur in every 2 to 3 year-interval, with cases increasing every year. In 2019, there were more than 14,000 cases with six reported deaths. In current outbreak of 2022, until October, there have already been around 34,000 cases with forty-four reported deaths.^8,9^

DENV belongs to the family Flaviviridae with four different serotypes (DENV-1, DENV-2, DENV-3, and DENV4). Infection by one serotype could confer long-term immunity to that serotype but subsequent infection by another serotype causes severe disease including DHF/DSS.^11-12^ Some studies have shown that DENV2 and DENV3 cause more severe illness than other serotypes.^13^ Therefore, DENV serotyping is important for disease management and public health surveillance.

Several methods can be used to diagnose of DENV infection and their utility depends on what a patient presents after symptom onset. However, due to limited availability of accurate diagnostics, many cases are also misdiagnosed, further underestimating the burden of disease.^14^ Virus isolation are technically challenging, expensive and require sophisticated biosafety.^15^ Serological methods, such as enzyme-linked immunosorbent assays (ELISA), may confirm the presence of a recent (IgM) or past infection (IgG) (through the detection of anti-dengue antibodies) but cannot identify the serotype and in some cases, the antibody can remain undetectable.^2,14^ Nucleic acid tests on the other hand, have relatively higher specificity and sensitivity and are also recommended by WHO.^16-17^ Additionally, molecular tests such as qPCR (quantitative Polymerase Chain Reaction or real-time PCR) are also capable for differential diagnosis of dengue, where there are other clinically indistinguishable infectious diseases such as malaria, leptospira and rickettsia.^18^ Some studies in Nepal have utlised Polymerase Chain Reaction (PCR) and sequencing to identify the circulating DENV serotype in previous outbreaks.^8,19-21^ However, regular molecular investigation is essential, when it has been reported that there are possibilities of replacement of one DENV serotype by another.^12,22^ For instance, in Sri Lanka, DENV3 was replaced by DENV1 in severe dengue epidemic of 2009.^12, 22, 23^

Nepal has faced periodic but major outbreaks in 2006, 2010, 2013, 2018 and 2019, there has been very limited molecular data on the circulating serotypes in the country (dominant serotypes: DENV-1/2, DENV-2, DENV-1, DENV-2, and DENV-2/3, respectively in 2010, 2013, 2016, 2017, and 2019). ^9,24-26^ Therefore, in this study, we have aimed to perform molecular investigation of DENV cases, of the ongoing outbreak, through the utilisation of multiplex-qPCR /Reverse Transcriptase PCR which will fill the information void and provide insights to the progression of dengue outbreaks.

## Methodology

### Samples Selection

In this study, we selected 100 dengue-positive serum samples, which were stored in -80C at Dhulikhel Hospital. The samples belonged to the dengue outbreak of 2022, collected between September-October 2022. They were, first, tested by serology and upon testing positive, were stored in the freezer.

### Dengue Serology

The serum samples were evaluated using Dengue NS1 Ag and IgM/IgG Ab Kit (Biotrol Laboratories). The samples were evaluated as dengue positive if they tested positive for either NS1 and/or IgM and/or IgG.

### Extraction and qPCR

Viral RNA was extracted from the serum samples using Zymo Quick RNA Viral Kit (Zymo, Cat: R1034-1034E). The extracted RNA (Ribo-Nucleic Acid) was evaluated using Sacace Dengue Real Genotype qPCR kit (Sacace, Cat: V63-50FRT) with use of proper controls. Negative process controls were used during extraction and PCR. The following master-mix recipe and thermocycling was used for serotyping:

After preparation, 15ul of the above master-mix was added to 10ul of extracted RNA.

### Data Interpretation

The kit detects presence of four serotypes through four separate channels in a single multiplex reaction as: DENV genotype 1 cDNA is detected in the FAM/Green channel; genotype 2 cDNA in the JOE/HEX/Yellow channel; genotype 3 in the Rox/Texas Red/Orange channel; genotype 4 in the Cy5/Red channel and Internal Control (IC) is detected in the Cy5.5/Crimson/ Quasar705 channel. As per the kit, the results were interpreted by the instance of crossing or not crossing of the threshold line by the fluorescence curve. The result of amplification was considered positive if the fluorescence curve was characteristic of sigmoidal curve and crossed the threshold line once in the significant fluorescence increase section and if the Ct value detected in the channel was below 38. Similarly, the result of amplification was considered negative if the fluorescence curve was not sigmoidal and if it did not cross the threshold line. The result was considered invalid if Ct value in the channel for the Cy5.5 fluorophore was not determined (was absent) or greater than the specified boundary Ct value.

## Results

### Samples

Out of 100 samples, only 89 dengue sero-positive samples were selected. The remaining 11 samples were excluded because either they did not have associated information or did not have sufficient volume.

### Subject Demography

Among the selected 89 subjects, 46 (52%) were female and the median age was 31 years (IQR, 7 to 68 years). Age wise positive cases of dengue was observed highest in age group between 21-30 years which constituted 40.4% of total cases whereas lowest was observed in age group above 51-60 and 61-70 years each of which comprised of 5.6% of the total cases. (Figure 1)

**Figure 1:**
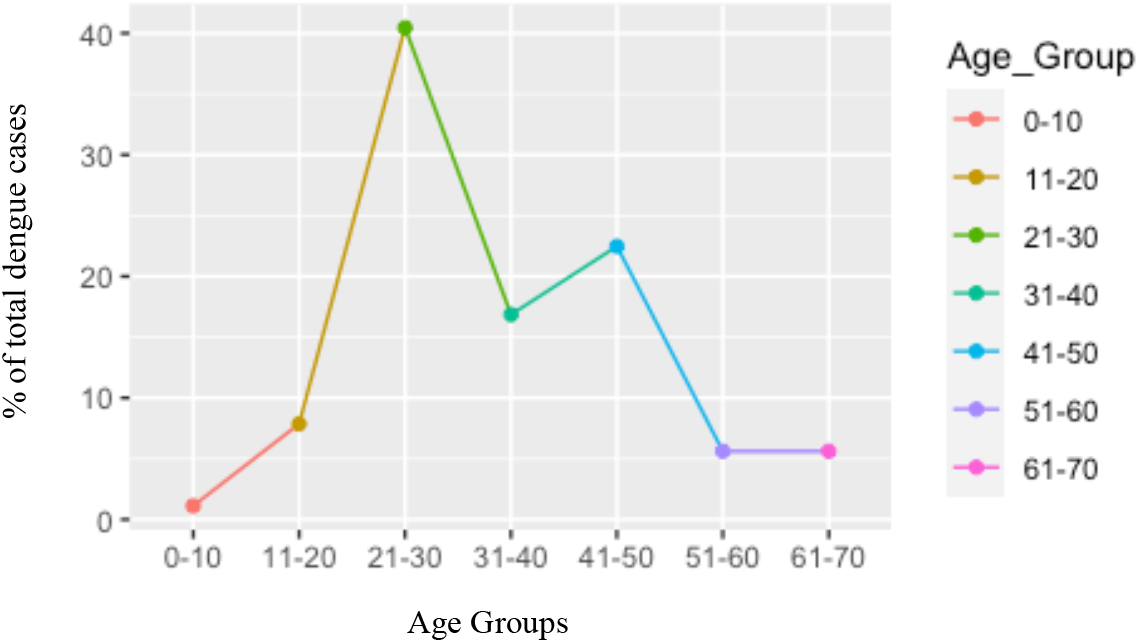
Distribution of sero-positive Dengue cases among age groups.

The participants were from different districts of Bagmati Province such as Kavre (54), Bhaktapur (9), Kathmandu (8), Sindhupalchowk (5), Sindhuli (3). The positivity was highest in Kavre District (58.4%). (Figure 2).

**Figure 2:**
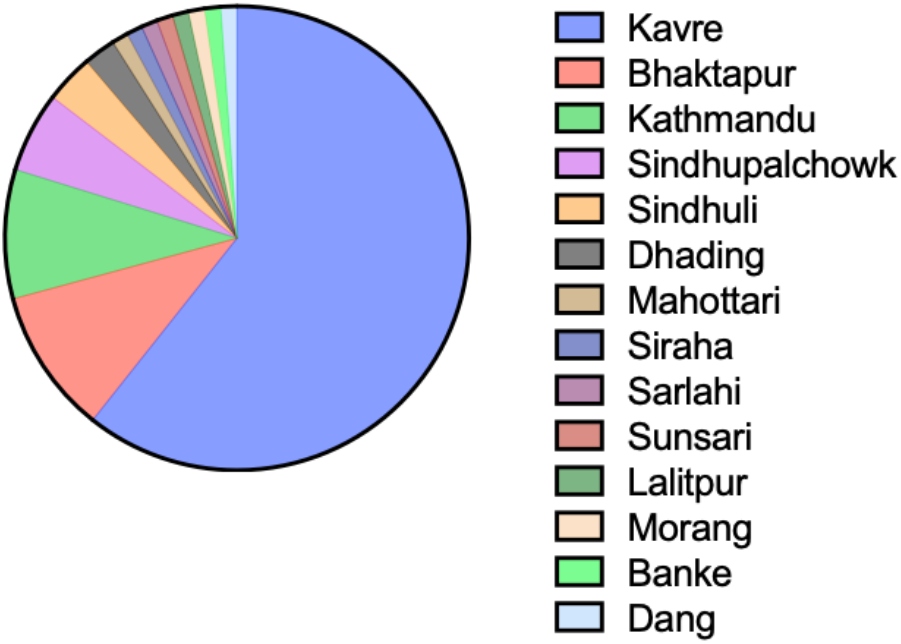
Distribution of sero-positive dengue cases by districts.

### Dengue Serology

Out of 89 samples, 76.4% (n=68) tested positive for NS1 only, 7.8% (n=7) tested for IgM only, while 2.2% (n=2) tested for IgG only. Similarly, 4.4% (n=4) tested positive for NS1 and IgM both, 2.2% (n=2) tested for NS1 and IgG both, 2.2% (n=2) tested for IgM and IgG both, while 4.4% (n=4) tested positive for all three.

### Dengue qPCR

The qPCR investigation confirmed the presence of 3 dengue serotypes: DENV1-3. Dengue serotype 4 was not detected in any sample. (Table 3)

**Table 1:**
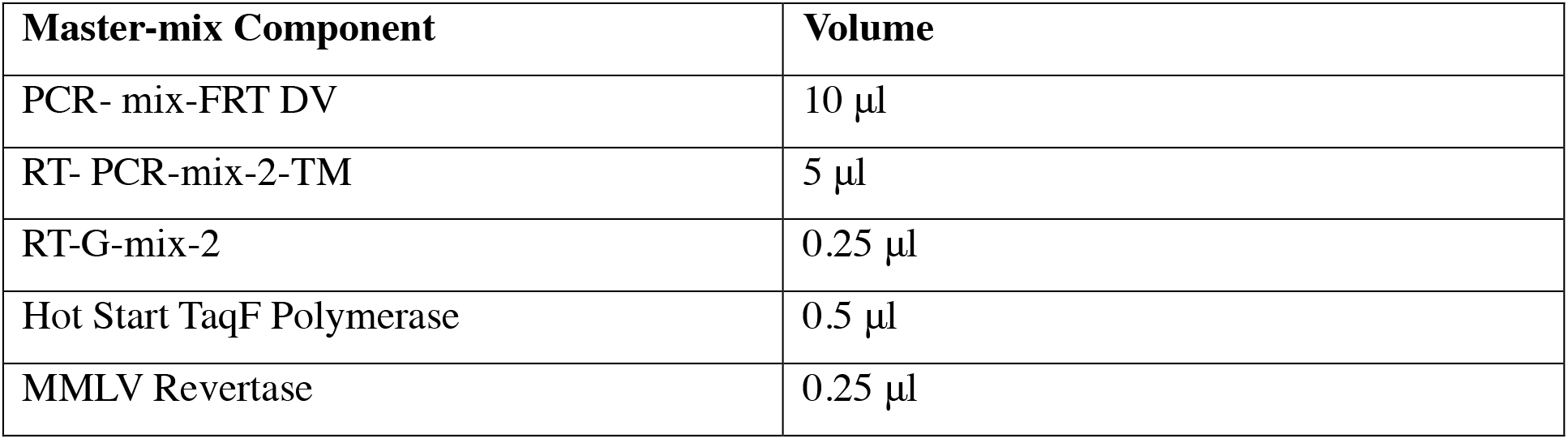
Master-mix Preparation for Sacace Dengue Real Genotype qPCR kit.

**Table 2:** Thermocycling parameters for qPCR.

| Temperature (°C) | Time | Cycles |
| --- | --- | --- |
| 50 | 30 mins | 1 |
| 95 | 15 mins | 1 |
| 95 | 10 secs | 5 |
| 56 | 40 secs |  |
| 72 | 20 secs |  |
| 95 | 10 secs | 40 |
| 54 | 40 secs (Fluorescence Capture) |  |
| 72 | 20 secs |  |

**Table 3:** Distribution of several DENV serotypes.

| SN | Serotypes | Number (%) |
| --- | --- | --- |
| 1 | DENV1 | 31 (58.4%) |
| 2 | DENV2 | 9 (16.9%) |
| 3 | DENV3 | 13 (24.5%) |
| 4 | DENV4 | N/A |

**Table 4:** Average Ct value, indicating subsequent viral load for each serotype.

| SN | Serotypes | Average Ct Value |
| --- | --- | --- |
| 1 | DENV1 | 18.07 |
| 2 | DENV2 | 17.3 |
| 3 | DENV3 | 16.6 |
| 4 | DENV4 | N/A |

The average Ct value of all serotypes was found to be 17.6, indicating high viral load. Additionally, the average Ct values were similar for different serotypes. The average Ct value for each serotype has been shown in the table below:

### Dengue Serology vs qPCR

When compared to prior serology, only 53 samples, out of 89, tested positive for Dengue by qPCR. From the comparison of 53 samples, it was observed that all (except one) of the sero-positive but qPCR negative samples were positive for either NS1 or IgM or both, while 27 NS1 sero-positive were negative by qPCR. (Figure 3)

**Figure 3:**
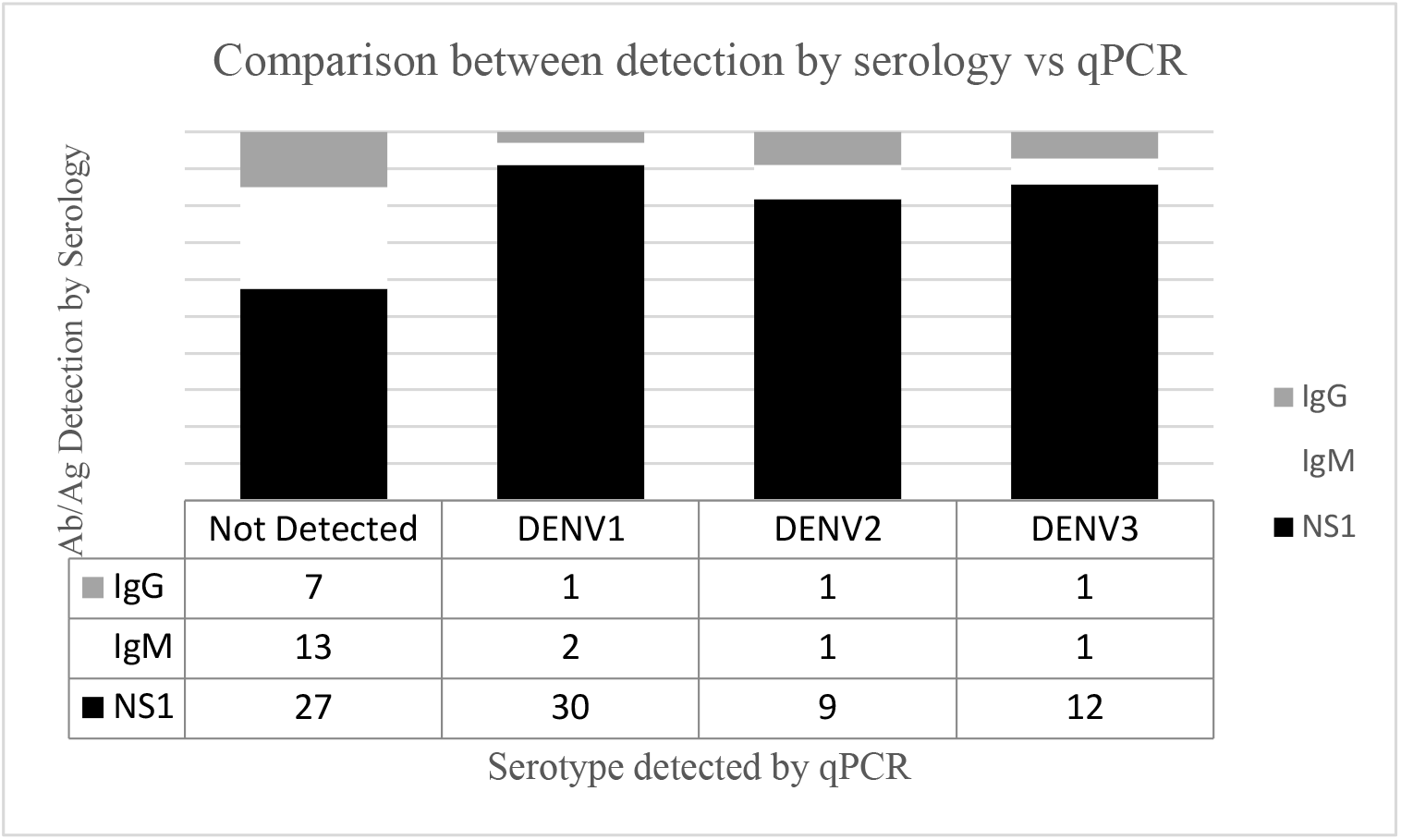
Comparison between dengue detection by serology (NS1, IgM and IgG) and qPCR (for each serotype) including not detected by qPCR.

## Discussion

### Subject Demography

Several studies have shown that risk of dengue fever and DHF varies by age.^27-30^ Though dengue is considered to be pediatric disease in most of South East Asia^31^, most adult patients present with dengue fever instead of DHF.^32^ Another study in Brazil also found that adults are more likely to have dengue compared to children.^33^ In our study, the median age of dengue positives was 31 years with 21-30 age group representing highest number of cases, while in other studies this has ranged from 6.8 to 24.3 years.^31,34,35^ Similar study in Nepal, however, showed adult median age (37 years).^36^ Interestingly, studies have shown that median age of infection, from dengue, has increased throughout the years.^34,36-38^ The increase in median age could be due to changes in surveillance practices, reduced mosquito-human contacts as well as shift in population demographics.^39^ There could also be change in age specific reporting and clinical presentation of dengue infection. Additionally, aging could have effects on the adaptive and cell mediated immunity leading to poor cytokine responses against dengue infection.^40^ Moreover, there could be other several factors such as virus serotype that could influence the correlation between age and prevalence of dengue.^27,33,41^

In our study, dengue positivity was found to be slightly higher in female. However, the correlation of dengue fever with gender have been variable in several studies. Some studies have shown higher prevalence among male^42-44^, while some have not found any correlation.^35^ However, the gender related correlation could change over time and could differ across countries based on different socio-cultural practices.^45^ Around our study site, female tend to work with household chores while male have tendency to work in nearer urban areas. This could possibly lead to exposure to environmental risk factors of dengue, in female.

In our study, the dengue-infected individuals were mostly from Kavre district which consists of sub-tropical and temperate regions.^46^ Traditionally, dengue infection has also been prevalent in tropical and sub-tropical regions, worldwide.^3^ However, the vectors can adapt to new environment and climate. The vectors for dengue have gained tolerance against colder climates, as an egg and adult.^47,48^ In Nepal, previously, in the first outbreak in 2006, only nine districts were affected by dengue, which increased to 45 districts in 2018 and 68 districts in 2019. These dengue cases, which were found at lower altitudes (lowland Terai), had spread to all of 77 districts in the outbreak of 2022, which now included high altitude regions.^8-10,49,50^ In 2019, Kavre district constituted of 0.3% of total the cases, which increased to 1% in 2022. Similarly, this percentage increased from 10.79% to 26.11% in Kathmandu.^51,52^ The reason behind this could be due to adaptation of mosquito responsible for dengue and increase in temperature in Nepal, resulting in ability to survive and breed even at altitudes of 2100m, which was not observed in prior years.^53^ One estimate suggested that high elevated mid hills have warmed up by 1.5 °C in the last 25 years, which is much higher than global average.^54^ Thus, there is higher risk of dengue expanding more northwards in the future.^26^

### Dengue serotyping

In this study, DENV1 was predominant, followed by DENV3 with absence of DENV4. There was no observable correlation in the serotype when compared to demography (age, gender) and geography. Unfortunately, the role of socio-demography is context specific in relation of DENV serotypes.^55^ Nevertheless, the overall distribution of serotypes, from this study, was similar to other studies in Nepal, done during epidemic of 2022, where DENV1 was observed to be highly prevalent followed by DENV3.^56-58^ Though Nepal has previously reported all four serotypes in the past, DENV1 and DENV2 have historically contributed to highest burden.^20,51^ In 2010 and 2016, DENV1 appeared while DENV2 was responsible for the outbreak in 2013.^59^ Similarly, only DENV1 and DENV2 were observed in outbreak of 2019.^51^ The re-emergence of DENV3 in 2022 is very alarming because the subsequent cross infection could lead to severe conditions of DHF and DSS.^11,12^

This study observed the average Ct value from qPCR to be 17.6. This low Ct value indicates that higher viremia which could propel the severity of dengue infection.^60^ Furthermore, the infection with DENV3 had slightly higher viremia levels (lower Ct value) compared to other serotypes. This could have been due to re-emergence of DENV3 after several years.^56,61,62^ But the difference was not significant enough to confirm this hypothesis.

### Dengue Serology vs qPCR

In this study, 69.5% of the serology positive serum samples were positive by qPCR. The discrepancy is unlikely due to procedural error because the Internal Control (during RNA extraction) and qPCR controls were all present, valid and in range with manufacturer manual.^63^

The difference in detection between the two methods is plausible as the sensitivity of the approaches vary with test principles and the time of sample collection during the infection period.^64^ Although the detection of viral RNA and NS1 antigens are generally suitable markers for DENV infection, NS1 antigen and IgM detection perform superior to RNA detection from day 6 to 7.^65^ Additionally, the time of sample collection from onset of fever is important because the antibody produced could lead to neutralization of virus, during late period of infection.^66^ This neutralization could result in lack of detection by qPCR. As our study included the retrospective samples, we could not identify the time of sample collection during the infection period. Another plausible explanation could be RNA degradation during storage and transport, causing lowered RNA concentration and negative result.^67^

Additionally, the observation of NS1 and IgM sero-positive but qPCR negative samples could indicate RNA degradation in samples.^68^ A similar study in Nepal also observed only one positive in qPCR out of 18 IgM positive samples.^69^ This study hypothesized degradation of viral particles, due to thawing, transportation and storage, as possible reason behind the discrepancy. Furthermore, the observation of qPCR negativity for two IgG only cases is plausible as IgG indicates past infection, implying that the samples may not have any viral particles (at the time of sample collection).^70,71,72^ Interestingly, there have also been studies that observed cross-reactivity between DENV and SARSCoV-2 antibodies.^73-76^ Such cross reactivity could have provided false positive in serological tests.

One of the limitations in this study was absence of information regarding clinical manifestation and thus, its association with DENV serotype could not be established.^75^ The incorporation of retrospective samples was the reason behind this limitation. Owing to the same reason, we could not identify the risk factors associated with DENV. However, our study was not designed to understand the associated risk factors. On the contrary, there are several strengths of this study. Firstly, the participants included individuals from young to old age group, which help address the epidemiological gap of age-specific serotype prevalence.^76,77^ Secondly, the population incorporated in the study was from outbreak, therefore represented general population visiting the hospital. Thirdly, the coverage of patients visiting the study site was diverse. The study site, Dhulikhel Hospital, hosts patients from wide range of geographical (terai and hilly region), social (mixed castes and ethnicity) and economic areas (urban and peri-urban).^78^ Nevertheless, this study was still able to overcame the gap posed by limited molecular data on circulating serotypes in Nepal.

## Conclusion

Dengue fever possess higher risk than ever with new cases emerging in higher altitude due to climate change. The risks posed in correlation to age and gender has been variable, with older age posing higher risk for dengue. Interestingly, there has been variation in detection of dengue through serology and qPCR, owing to the difference in sensitivity, time of sample collection, RNA degradation or cross-reactivity.

The current outbreak showed highest prevalence of DENV1 followed by DENV3. The re-emergence of DENV3, with high viral load, compared to preceding outbreak of 2019 could cause cross-infection leading to severe conditions of dengue hemorrhagic fever/dengue shock syndrome. Thus, the findings from this study, of dengue outbreak of 2022, was able to fill the void due to limited molecular data on the circulating serotypes.

## Data Availability

All data produced in the present study are available upon reasonable request to the authors.

## Acknowledgements

We sincerely thank Nepal Health Research Council for funding this study through Provincial Health Research Grant 2079/80. We appreciate Department of Microbiology, Dhulikhel Hospital for providing the samples and related information.

